# The Basel Long COVID Cohort Study (BALCoS): protocol of a prospective cohort study

**DOI:** 10.1101/2024.10.29.24316282

**Authors:** Stefan Rohner, Rebekka Schnepper, Gunther Meinlschmidt, Rainer Schaefert, Michael Mayr, Katrin Bopp, Andrea Meienberg

**Affiliations:** University of Basel, Faculty of Medicine, Basel, Switzerland; University Hospital Basel, Department of Psychosomatic Medicine, Basel, Switzerland; University Hospital Basel, Department of Digital and Blended Psychosomatics and Psychotherapy, Basel, Switzerland; University of Trier, Department of Clinical Psychology and Psychotherapy, Trier, Rheinland-Pfalz, Germany; University Hospital Basel, Medical Outpatient Clinic, Basel, Switzerland

**Keywords:** cohort study, SPIROS, post-COVID, long COVID, post-acute sequelae of SARS-CoV-2, PASC, study protocol

## Abstract

**Introduction:** The recent severe acute respiratory syndrome coronavirus type 2 (SARS-CoV-2) pandemic had a devasting global impact. Many people suffered from coronavirus disease 2019 (COVID-19) and some experienced persistent symptoms interrupting their lives even further. The World Health Organization (WHO) defined the condition of these persistent symptoms as post-COVID-19 condition (PCC). The most prevalent PCC symptoms are fatigue, dyspnea, sleep disturbances, coughing, anosmia and ageusia, chest pain, and headaches. This article describes the protocol of the Basel Long COVID Cohort Study (BALCoS), which aims at fostering understanding of PCC and investigating underlying mechanisms for the development and course of the condition by focusing on participants’ health status and symptoms with repeated measures over one year.

**Methods and analysis:** BALCoS is a prospective single site cohort study. Inclusion criteria are a confirmed PCC diagnosis according to WHO or a subjective attribution of persistent symptoms to PCC, proficiency in German to follow study procedures, and at least 18 years of age. It comprises blood sample collections, standardized neurocognitive and psychometric tests, physical performance measures, and ecological momentary assessments (EMAs). Standardized tests and EMAs are administered at baseline (BL), and at 3-, 6-, and 12-months follow-up. At BL and 12-month follow-up, physical performance and neurocognitive abilities are assessed. Participants provide blood samples at BL. The study is exploratory in nature and a sample size of at least 120 participants is targeted. The study is part of a larger Horizon Europe Long COVID project combining mechanistic, clinical, and intervention studies within an interdisciplinary European research consortium.

**Ethics and dissemination:** The Ethics Commission of Northwest and Central Switzerland approved the study (BASEC-ID: 2023-00359), which is registered at ClinicalTrial.gov (ID: NCT05781893). All participants provide written informed consent. Key results from the study will be published in peer-reviewed journals.

**Funding Details:** BALCoS is primarily funded by the Swiss State Secretariat for Education, Research and Innovation (SERI) under contract number 22.00094 in the context of the European Union’s Horizon Europe research and innovation program under grant agreement No. 101057553.

## INTRODUCTION

In early 2020, the coronavirus disease 2019 (COVID-19), caused by the severe acute respiratory syndrome coronavirus 2 (SARS-CoV-2), has led to a global pandemic. As the pandemic progressed, it became apparent that the disease not only causes acute symptoms but can also lead to long-term complaints (sometimes termed “long COVID”, in the following referred to as “post-COVID-19 condition”, PCC). According to the definition of the World Health Organization (WHO), PCC is defined as symptoms occurring in individuals with a history of confirmed or suspected SARS-CoV-2 infection, lasting for at least two months, usually within three months from the onset of the acute infection, that cannot be explained by an alternative diagnosis (1). Generally, symptoms are heterogeneous (2) and present across multiple organ systems. Common symptoms include fatigue, dyspnea, sleep disturbances, coughing, anosmia and ageusia, chest pain, headaches (3). According to a meta-analysis conducted in March 2022, the prevalence of PCC ranges from 7.5% to 41% in non-hospitalized adults and averages 37.6% in hospitalized adults (4). Due to methodological differences in the published literature, prevalence rates vary considerably and numbers are difficult to reliably estimate. Even though some risk factors of PCC could be identified, such as virus variant, female gender, hypertension, smoking, obesity, and comorbid diseases (5-9), the etiology of PCC is still unclear (10, 11). Several hypotheses have been proposed, such as immune dysregulation (12), microbiota dysbiosis which affects the microbiota and virome, including the persistence of SARS-CoV-2 (13), autoimmunity and primed immune cells by molecular mimicry (14), microvascular coagulation with endothelial dysfunction (15), and dysfunctional signaling in the brainstem and/or vagus nerve (16). These findings show that biomedical components have been the focus of PCC research. However, psychological factors are also increasingly considered and have been shown to play an important role (17). These factors are incorporated into psychotherapeutic treatment approaches (18). Due to insufficient understanding of the mechanisms that underlie the development of PCC, treatment options are currently limited and mainly focus on symptom management. This lack of understanding potentially contributes to those affected by PCC experiencing feelings of helplessness, frustration, being misunderstood, worry due to the uncertain course of the disease, and a sense of losing control over their lives (19). A better understanding of the pathogenesis, risks, and protective factors of PCC would help to improve health care and support of people affected with PCC (20, 21). Here we provide the protocol of the Basel Long COVID Cohort study (BALCoS), which aims to investigate and better understand PCC, its symptoms and course, and to illuminate the underlying multifactorial mechanisms of the condition by collecting a broad biopsychosocial spectrum of data over the course of one year. Participants complete a set of assessments that include the collection of sociodemographic and clinical data, biomarkers, neurocognitive testing, psychometric questionnaires, ecological momentary assessments (EMAs), and measures of physical performance. This research serves as a basis for planning future measures and provides information on clinically relevant factors for health care professionals. From this research it may be possible to derive factors that positively influence the quality of care of patients and their overall quality of life.

## METHODS AND ANALYSIS

### Study Design and Setting

BALCoS is a registry-based, single-center, prospective, observational cohort study. The study site is located at the Medical Outpatient Clinic of the University Hospital Basel (UHB) and participants are recruited from patients attending the specialized PCC consultation service at the UHB. In addition, study participation is open to individuals from consultation services and clinics outside the UHB and individuals who respond to study advertisements. Written consent is required from all participants.

### Study Timeline

Recruitment started in February 2023, with the first participant enrolled in April 2023. The study is intended to extend beyond the current SERI-funded research period, which runs until May 2026 to allow for further data collection and analysis, as well as continued scrutiny of the research questions, including changes in PCC over time (see Figure 1).

**Figure 1.**
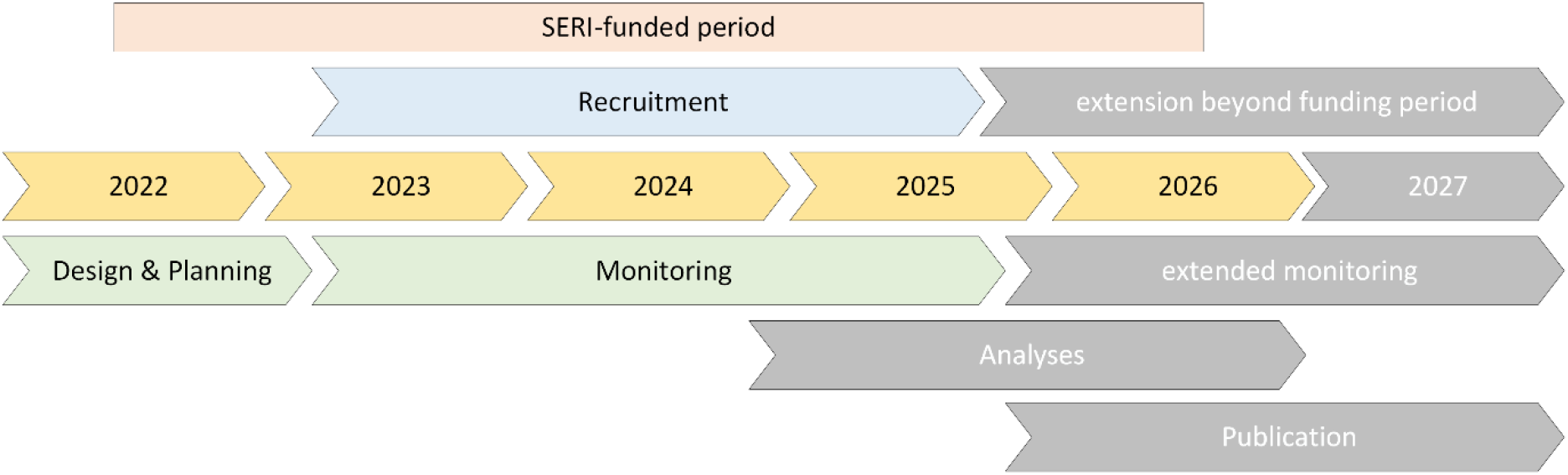
Timeline of the Basel Long COVID Cohort Study. Note: SERI = State Secretariat for Education, Research, and Innovation; COVID = coronavirus disease.

### Study Schedule

Participants are asked to give informed consent to the inclusion of all their data from previous medical examinations that could be relevant to their PCC symptoms. Further, they are asked to agree that the data collected in the study can be used for routine clinical care. Participants not recruited from the UHB PCC clinic provide this information in an enrollment interview, after signing the informed consent form. An overview of all clinical information collected at the baseline interview and routine clinical examination can be young in the supplemental material. At baseline (BL) and T3, participants complete psychometric measures and neurocognitive tests on a computer, tablet, or similar device. If the BL assessment is too exhausting for one visit, a second visit can be arranged to complete the neurocognitive test battery and measures of physical performance (marked with “(x)” in Table 1). Participants can choose to complete the study remotely, if travelling to the study site is not feasible due to health concerns or symptom burden. These participants complete the study as planned, except for the completion of physical performance measures, neurocognitive assessments, and blood sample collection at BL. Remote participants receive a link by email to complete the psychometric measures for BL and T3 online at home. At T1 and T2, all participants (remote and in-person) receive a link by email to a battery of psychometric questionnaires to complete online at home. One day after completing each data collection point (BL, T1, T2, and T3), participants receive a link via text message at 6 p.m. directing them to the EMA questions. These 8 EMA questions are answered over ten consecutive days. A detailed summary of the study plan is provided in Table 1.

**Table 1.** Outcome measures and assessment schedule.

|  | Outcome measure | BL |  | T1 | T2 | T3 |
| --- | --- | --- | --- | --- | --- | --- |
|  |  | 1 | 2 |  |  |  |
| 0 | Oral and written information, written consent, screening of inclusion and exclusion criteria | x |  |  |  |  |
| 1 | Blood samples* | x |  |  |  |  |
| 2 | Neurocognitive assessment: Central Nervous System Vital Signs (CNSVS) * | x | (x) |  |  | x |
| 3 | Physical performance measures* |  |  |  |  |  |
| a | 6-minute walking test | x | (x) |  |  | x |
| b | Jamar dynamometer | x | (x) |  |  | x |
| c | 1-minute sit-to-stand test (STS-60) | x | (x) |  |  | x |
| 4 | Psychometric questionnaires |  |  |  |  |  |
| a | Functional Capacity: World Health Organization Disability Assessment Schedule (WHODAS 2.0) | x |  | x | x | x |
| b | Visual Analogue Scales (VASs): quality of life, symptom intensity, functional impairment, work ability | x |  | x | x | x |
| c | Quality of Life: Quality-of-Life-8 (EuroHIS QOL-8) | x |  | x | x | x |
| d | Somatic Symptom Severity: Patient Health Questionnaire (PHQ-15) | x |  | x | x | x |
| e | Somatic Distress: Somatic Symptom Disorder – B Criteria Scale (SSD-12) | x |  | x | x | x |
| f | Depression : Patient Health Questionnaire Depression Scale (PHQ-8) | x |  | x | x | x |
| g | Anxiety: Generalized Anxiety Disorder Questionnaire (GAD-7) | x |  | x | x | x |
| h | Resilience: Resilience Scale (RS-11) | x |  | x | x | x |
| i | Insomnia: Insomnia Severity Index (ISI) | x |  | x | x | x |
| j | Fatigue : Chalder Fatigue Scale (CFS) | x |  | x | x | x |
| k | Stress: Perceived Stress Scale (PSS-10) | x |  | x | x | x |
| 6 | Ecological momentary assessments (EMAs; 8 daily questions for 10 consecutive days after BL, T1, T2, T3) |  | x | x | x | x |

Note: BL = Baseline
\*Measurements not completed by remote participants

### Sampling procedure and selection

Enrollment is based on the inclusion criteria (see Table 2) and recruitment is carried out by the study team which includes physicians, psychologists, psychotherapists, and study nurses.

**Table 2.** Inclusion and exclusion criteria.

| Inclusion criteria | Exclusion criteria |
| --- | --- |
| Confirmed PCC diagnosis according to WHO or subjective attribution of persistent symptoms to PCC | Lack of general understanding of the study procedures |
| At least 18 years of age | Rejection of consent to participate |
| Adequate knowledge of German |  |

Note: PCC = post-COVID-19 condition

Until June 2024, only patients attending the PCC clinic at UHB were recruited. Due to recruitment difficulties, recruitment was expanded from July 2024 to include patients from other specialized consultation services or clinics in the broader Basel area with physician-based diagnosis. From September 2024 onward, individuals with PCC symptoms and subjective attribution of their persistent symptoms to PCC have been recruited as well. The study is advertised in other specialized and outpatient clinics, general practitioners’ offices, on public transportation, at conferences and symposia, mailing lists, and via social and other digital media. All potential participants are screened for inclusion and exclusion criteria. The study investigator (or their nominee) obtains written informed consent from all study participants. Study information is presented by phone or video call if travel for the sole purpose of face-to-face recruitment is not possible. As part of the recruitment and informed consent process, the investigators (or their designees) explain to each participant the nature of the study, its purpose, the procedures involved, the expected duration, the potential risks and benefits, and any inconvenience that may be involved. Each participant is informed that participation in the study is voluntary, that they may withdraw from the study at any time, and that withdrawal of consent will not affect their subsequent medical care. No formal sample size calculation and or power analysis has been performed prior to recruitment. The target sample size is 120 participants for the funded period. The rationale for no prior sample size calculation is based on the exploratory nature of the study. The target sample size is based on heuristics and estimates by the study team regarding of the number of people that sought help at the PCC clinic in 2021 and 2022.

### Horizon Europe Long COVID consortium

The BALCoS study is part of the Horizon Europe Long COVID project. The consortium takes a biopsychosocial perspective on PCC and aims to gain a more comprehensive understanding of PCC to identify mechanisms, processes, and biomarkers of the condition, and to develop management strategies and treatment approaches. The research efforts include cohort studies, biomechanistic studies, and (digital) intervention studies. BALCoS is one of the cohort studies being conducted. Participants in the BALCoS cohort are asked to provide blood samples for the consortium’s biomechanistic studies.

### Blood samples and assessments^1^

#### Blood Samples

At BL, participants provide blood samples for on-site analysis and for analysis by the consortium (see Table 3). These samples will be analyzed for genomic and human leukocyte antigen (HLA) typing (plasma), COVID-19 antibody profiling (serum), lipidome (serum), autoantibody epitopes (serum), and coagulation analyses (citrated plasma). Due to limited resources, for some parameters only a sub-sample of enrolled participants will be sent and analyzed. Participants may refuse genomic analyses and still be included in the study.

**Table 3.** Overview of blood sample collection.

| Blood samples for on-site analysis | Blood samples for the consortium |
| --- | --- |
| RNA tubes for genetic analyses (optional) | EDTA plasma (for genetic analyses; optional) |
| EDTA plasma | Citrated plasma |
| Serum | Serum |

Note: RNA = Ribonucleic Acid, EDTA = Ethylenediaminetetraacetic Acid.

#### Neurocognitive Testing

*CNS Vital Signs* (CNSVS, 22) is a computerized neurocognitive test battery that was developed as a routine clinical screening instrument. It consists of seven scientifically valid and reliable neuropsychological tests. The tests are verbal and visual memory, finger tapping, symbol digit coding, the Stroop Test, a test of shifting attention, and the continuous performance test. Completion of the tests takes 40-50 minutes. The psychometric properties of the CNSVS tests have been shown to be comparable to the interview-based conventional neuropsychological tests from which they were derived (23).

#### Psychometric Questionnaires

*Visual Analogue Scales (VAS, 24) –* Participants answer 4 questions on a VAS with a 10-point Likert scale rating slider ranging from 1 to 10. The questions cover symptom intensity (“How do you currently rate the intensity of your symptoms?”, 10 = it can’t be worse), functional impairment (“How do you currently rate the impairment caused by your symptoms in everyday life?”, 10 = it can’t be worse), quality of life (“How would you rate your current quality of life?”, 10 = it can’t be better), and work capacity (10 = full work capacity). For work capacity, participants indicate their current work capacity (“current workload in % per week”) and their maximum work capacity before the onset of PCC symptoms on a scale from 0% - 100% (“Regular workload before symptoms started in % per week”).

*Functional Capacity* – World Health Organization Disability Assessment Schedule (WHODAS 2.0): The WHODAS 2.0 assesses and classifies disability due to health problems during the 4 previous weeks. The 12-item version is used in this study (25). An example item would be “In the past 4 weeks, how much difficulty have you had walking a long distance such as one kilometer (or equivalent)?”. Items are answered on a 5-point Likert scale ranging from 0 = “none” to 4 = “extreme or impossible”.

*Quality of Life* – EuroHIS Quality-of-Life-8 (QOL-8, 26): This 8-item instrument assesses quality of life and perceived health during the 4 previous weeks, e.g. by asking “Do you have enough energy to cope with everyday life?”. Items are answered on a 5-point Likert scale, with wording varying between questions (e.g., from “very dissatisfied” to “very satisfied”).

*Somatic Symptom Severity* – Patient Health Questionnaire (PHQ-15, 27): The PHQ-15 assesses the severity of somatic symptom over the previous 4 weeks. Participants are asked how much they felt affected by their symptoms, (e.g., back pain). Items are answered on a 3-point Likert scale ranging from 0 = “not bothered at all” to 2 = “bothered a lot”.

*Somatic Distress:* Somatic Symptom Disorder–B Criteria Scale (SSD-12, 28):The SSD-12 assesses psychological features of somatic disorders with four questions each on the cognitive (disproportionate and persistent thoughts about the seriousness of one’s symptoms), affective (health anxiety), and behavioral (excessive time and energy devoted to these symptoms or health concerns) dimensions. Items are answered on a 5-point Likert scale ranging from 0 = “never” to 4 = “very often”.

*Depression* – Patient Health Questionnaire Depression Scale (PHQ-8, 29): The PHQ-8 assesses the severity of depressive symptoms according to the 4^th^ Edition of the Diagnostic and Statistical Manual of Mental Disorders (DSM-IV;30). Participants are asked whether they have been bothered by symptoms typical for depression (e.g., by having little interest or pleasure in doing things) in the past few weeks. Items are answered on a 4-point Likert scale ranging from 0 = “not at all” to 3 = “almost every day”.

*Anxiety* – Generalized Anxiety Disorder Questionnaire (GAD-7, 31): The GAD-7 measures anxiety. It assesses whether the participant has been bothered by complaints related to anxiety in the past 2 weeks (e.g., by asking about difficulties in relaxing). Items are answered on a 4-point Likert scale ranging from 0 = “not at all” to 3 = “almost every day”.

*Resilience* – 11-item Resilience Scale (RS-11, 32): The RS-11 asks about general resilience to life events, e.g. “I feel that I can handle many things at the same time.”. Items are answered on a 7-point Likert scale ranging from 1 = “strongly agree” to 7 = “strongly disagree”.

*Insomnia* – Insomnia Severity Index (ISI, 33): The ISI assesses sleep problems. Items are answered on a 5-point Likert scale regarding severity over the past 2 weeks, with wording varying between questions (e.g., from 0 = “very dissatisfied” to 4 = “very satisfied”).

*Fatigue* – 11-item Chalder Fatigue Scale (CFS, 34): The CFS assesses fatigue on a mental subscale (e.g., by asking “Do you have difficulty concentrating?” and a physical subscale (e.g., by asking “Do you need to rest more?”). Items are answered on a 4-point Likert scale ranging from 0 = “better than usual” to 3 = “much worse than usual”.

*Stress* – Perceived Stress Scale (PSS-10, 35): The PSS assesses the experience of psychological stress in the last 4 weeks. It consists of 10 items, e.g., “In the last month, how often have you been upset about something that happened unexpectedly?”. Items are answered on a 5-point Likert scale ranging from 1 = “never” to 5 = “very often”.

*EMA Questions* – After each data collection point (BL, T1, T2, T3), participants answer 8 daily EMA questions for 10 consecutive days. Questions 1-5 are based on the Screening Tool for Psychological Distress (STOP-D, 36), question 6 is based on the EQ-5D (37, 38), question 7 is based on the Single Item Mindfulness Scale (SIMS; 39), and question 8 is based on the Single Item Self Compassion Scale (SISC; 40). The questions are completed in the Research Electronic Data Capture (REDCap, 41) on a 10-point Likert scale ranging from 0 = “not at all” to 9 = “severely”. Namely, these questions are: “*In* the last 24h, how much have you been…

1. …bothered by feeling sad, down, or uninterested in life?
2. …bothered by feeling anxious or nervous?
3. …bothered by feeling stressed?
4. …bothered by feeling angry?
5. …bothered by not having the social support you feel you need?
6. …bothered by difficulties in carrying out your daily activities?
7. …fully in the moment, accepting it as it is?
8. …high levels of self-compassion for yourself?”

#### Measures of physical performance

*Grip strength* – Jamar dynamometer (42): Sitting in a chair, the participant is instructed to squeeze the hydraulic grip strength measuring device with one hand, while the other arm rests in their lap. The test is repeated three times for each hand, first for the dominant hand, then for the non-dominant hand. The best value (in kg) for each hand is recorded. Injuries to one or both hands are also recorded.

*Functional exercise capacity* – 6-minute walking test (43): The distance (total meters), that a participant can walk in six minutes is measured. Pulse and oxygen saturation are assessed at baseline, immediately after finishing the test, and 2 minutes after finishing the test, using a pulse oximeter. Pre- and post-exercise comfort is rated on a 5-point Likert scale ranging from 1 = “very poor” to 5 = “very good”. After the test, perceived respiratory and leg exertion is assessed using the Borg Category-Ratio (CR) 10 Scale ranging from 0 = “no exertion” to 10 = “maximum exertion” (44).

*Muscular endurance* – Sit-to-Stand Test (STS-60, 45): The STS-60 assesses the number of sit-to-stand cycles that the participant can complete in 60 seconds without using their hands for support. Oxygen saturation at the end of the test is assessed using a pulse oximeter.

### Data collection and management

Data are mainly collected through electronic means: Psychometric questionnaires, EMAs and neurocognitive assessments in the form of online surveys or computerized assessments. Data collection for physical performance measures is paper based. Electronic data capture is done within the REDCap electronic case report form (eCRF). Paper-based data will be entered into REDCap by trained members of the study team. Clinical data will be retrieved from the patient’s medical records and merged into REDCap or collected in a clinical entry interview with external patients conducted by the study team and then entered into REDCap. Data quality is supported by continuous monitoring, plausibility checks of the collected data and checks for missing values by data managers. In addition, participants receive automated reminders from REDCap to complete the questionnaires at T1 and T2, three and seven days after the initial invitation. If the questionnaires remain incomplete, the study team contacts participants by phone two weeks after the initial invitation. The study team also monitors adherence to the EMA questions manually and contacts participants by phone if two consecutive days remain incomplete. Data management and monitoring is performed by authorized and experienced study investigators and project employees. In accordance with local laws and regulations, health-related data and biological materials are stored for ten years after the publication of the research project. Only authorized personnel have direct access to participant-level data and all electronic documents are stored on UHB-maintained and regularly backed-up server. As soon as possible after data collection all study-related documents are encoded, and participants are only identified by their unique participant number. Personal data (i.e., phone numbers and e-mail addresses) will be recorded in REDCap for the purpose of sending out the psychometric questionnaires and EMAs at T1 and T2. Any study-related or participants’ personal data are only accessible for authorized personnel (e.g., paper-based documentations are locked away in a research office). Members of the study team conducting any research activity (e.g., data monitoring or using a specific software) have been trained by senior members of the team and/or have completed a training course and are acting on Standard Operating Procedures (SOPs) set by UHB or developed by the team according to Good Clinical Practice (GCP).

### Potential Biases

The inclusion of external participants requires a subjective attribution of persistent symptoms to PCC or a confirmed PCC diagnosis. The diagnostic process of these instances may differ from the diagnostic process of the specialized clinic in the UHB, potentially introducing more heterogeneity into the sample. To address this potentially confounding variable, we will conduct independent t-tests to investigate whether the three groups (subjective attribution, medical diagnosis made outside or from the physicians from the UHB) differ in terms of sociodemographic and diagnostic parameters. Since participation in the study is voluntary, patients with high motivation, prosocial behavior, or mild or moderate PCC severity may introduce a self-selection bias into the sample, as patients with more severe PCC may not have the resources or energy to participate in the study, thereby potentially compromising the generalizability of the results.

### Statistical Analysis

Data will be analyzed in RStudio (46). Categorical data (e.g., sex) will be reported as counts and percentages. Metric data (e.g., age) will be reported as means with standard deviations. Primary analyses of longitudinal data include the calculation of generalized linear mixed models (GLMMs) for psychometric, neurocognitive, and physical assessment data, and the calculation of generalized additive mixed models (GAMMs) for EMAs. In both cases, participants are included as a random effect and time as a fixed effect. A priori subgroups of interest regarding interactions or effect modifying factors are sex, age, and duration of PCC symptoms at BL. Additional covariates will be considered if suspected of confounding influence. Secondary analyses include correlation analyses between EMAs and psychometric questionnaires.

Missing data will be addressed using appropriate statistical methods, such as multiple imputation, and is categorized into three groups: drop-out, lost-to-follow-up, and discontinuation. Drop-out is defined as clear, participant-initiated, written, or verbal withdrawal of consent before they complete the last data collection point (T3), in which case recorded participant data is deleted upon request. Participants opting to drop out of the study cannot reenter at a later point. Participants are discontinued from the study if they are not reachable and/or refrain from communicating with the study team before any data collection. In case any data was collected (e.g., blood samples from BL), they are declared lost-to-follow-up. Missing data from discontinued participants will not be imputed. Participants are contacted at least four times before they are informed that the study team is no longer trying to contact them to schedule study-related appointments or remind them of adhering to study procedures. Participants are firstly contacted by phone (with a message on the answering machine if the call is not answered), secondly by mail, thirdly via phone again, and then, lastly, by an email explaining that there will be no further contacting attempts. Depending on whether any data were collected, the participants are then either declared lost-to-follow-up or discontinued from the study.

## ETHICS AND DISSEMINATION

The study has been approved by the Department of Clinical Research at University Hospital Basel (ID: th22schaefert), the Ethics Commission of Northwest and Central Switzerland (ID: 2023-00359) and is registered at ClinicalTrial.gov (ID: NCT05781893). The first amendment to this study protocol was submitted to the local ethics committee March 18, 2024, and accepted March 22, 2024. The second amendment was submitted on June 3, 2024, and accepted June 18, 2024. Further amendments will be submitted to the appropriate bodies if needed. Key results from this study will be published in peer-reviewed journals. The full protocol is accessible through this manuscript. Study results are planned to be presented at conferences, symposia, lectures, and presentations to the scientific community and the public. Dissemination of results is additionally aligned within the consortium to optimize impact. Authorship eligibility of this publication is based on the International Committee of Medical Journal Editors (ICMJE) guidelines (47).

### Risks and Harm to Participants

There is no additional effort except for additional blood samples and detailed data collection, which requires cognitive and temporal efforts for the participants. Participants receive 50 Swiss Francs as compensation for taking part in the study after completing T3, with a proportional amount being paid out in case of withdrawal from the study. No intervention besides standard care including individual remedies is given, no radiation is used, and no vulnerable group is recruited. According to Human Research Ordinance (HRO) Art. 7 the risk category of this study is “A”. Adverse events and serious adverse events are reported according to HRO Art. 21 and the appropriate bodies to be notified of safety and protective measures according to the Human Research Act (HRA) Art. 15 and HRO Art. 20.

### Public and Patient Involvement

The measurements and data collection instruments were chosen to adequately reflect the range of PCC symptoms as presented by patients across the globe and informed by the newest published research at the time of study development. The experiences and feedback from patients treated in the PCC clinic at the UHB to date were taken into account when developing and planning the study.

### Artificial Intelligence

One author used Grammarly (48) for spell-checking this manuscript and two authors used DeepL (49) to help translate study materials to English.

### Guidelines

We used the Standardized Protocol Items Recommendations for Observational Studies (SPIROS, 50) in the preparation of this protocol, and we will use the Strengthening the Reporting of Observational Studies in Epidemiology (STROBE, 51) guidelines when reporting the results of the study.

### Open Science

There are currently no plans to grant public access to participant level data of this study. Participant level data and pooled study data will be made available for consortium partners as part of a joint controllership agreement. A copy of the informed consent form is available upon request. The statistical codes will be made available upon request after analyses are concluded.

## Supporting information

Routine clinical information

## Acknowledgements

The authors acknowledge the dedicated engagement and support of Julia Menges, Tabea Rocco, Nadine Celine Grimm, Michèle Häner, and Lukas Ebner. Their feedback on the study materials, inputs for planning and help in achieving core tasks in the study has been an invaluable contribution. Further, the authors acknowledge the support of the master students Aigerim Abdrakhim, Saskia Erni, Stefanie Meier, and Janik Hendry in the ongoing recruitment of participants and their overall contribution to the study. The authors would like to extend their gratitude to the doctors and general staff of the specialized PCC clinic for their efforts and contributions towards the success of the study.

## Author Contribution Statement

Contributions to this publication are structured according to the Contributor Roles Taxonomy (CRediT;52). All authors have contributed, read, and approved the final version of the manuscript.

Conceptualization: SR, ReS, GM, RaS, MM, KB, AM

Methodology: SR, ReS, GM, RaS, MM, KB, AM

Software: SR, ReS

Resources: SR, ReS, GM, RaS, MM, KB, AM

Writing - Original Draft: SR

Writing - Review and Editing: SR, ReS, GM, RaS, MM, KB, AM

Supervision: GM, RaS

Project administration: SR, ReS

Funding acquisition: GM, RaS, MM, KB, AM

## Strengths and Limitations

- Repeated assessments are expected to inform about the course of illness progression and development within routine medical care.
- A broad array of participant-reported outcomes strengthens participant perspective of symptoms.
- The duration of data collection and the number of questionnaires might influence questionnaire completion and adherence.
- Planned follow-up of 12 months limits conclusions regarding long-term outcome.
- The exploratory nature of the study allows to explore risk factors and associations, but not causality.

## Footnotes

1 All study materials including (computerized) measurements are in German only.

