## Supplementary material for "The Basel Long COVID Cohort Study (BALCoS): protocol of a prospective cohort study": Routine clinical information

### Supplemental Material : Routine clinical information

**Clinical Information**

Age (in years):

Sex:

| | | | |
|---|---|---|---|
| Male | Female | Diverse | Unknown/other |

Nationality:

| | | | |
|---|---|---|---|
| Swiss | German | French | Other – Please specify: |

Date of first clinic consulation:

Study inclusion at first clinic consultation:

| | | |
|---|---|---|
| Yes | Yes, with reservations | No |

**Assessment of Therapies**

<table>
<tr><th>Which therapies regarding PCC were used or are currently being used?</th><th>Answer Categories</th></tr>
<tr><td>Physical Therapy</td><td rowspan="27">
<ul>
<li>Therapy: Yes/No</li>
<li>Time frame:
<ul><li>Start date:</li><li>Currently in use: Yes/No</li><li>End date</li></ul></li>
<li>Frequency:
<ul><li>Daily</li><li>One or multiple times per week</li><li>At least once a month</li><li>Less than once a month</li></ul></li>
<li>Change of symptoms as part of therapy:
<ul><li>Clear improvement</li><li>Moderate improvement</li><li>Mild improvement</li><li>No change</li><li>Mildly worsened</li><li>Moderately worsened</li><li>Clearly worsened</li></ul></li>
<li>Additional specifics:</li>
</ul>
</td></tr>
<tr><td>Ergotherapy/occupational therapy</td></tr>
<tr><td>Acupuncture</td></tr>
<tr><td>Psychotherapy (individual setting)</td></tr>
<tr><td>Psychotherapy (group setting)</td></tr>
<tr><td>Non-drug studies</td></tr>
<tr><td>Self-help courses</td></tr>
<tr><td>Outpatient rehabilitation</td></tr>
<tr><td>Antihistamines</td></tr>
<tr><td>Apheresis</td></tr>
<tr><td>Hyperbaric oxygen therapy</td></tr>
<tr><td>Intermittend hypoxia-/hyperoxia therapy</td></tr>
<tr><td>Selective serotonin reuptake inhibitors (SSRI)</td></tr>
<tr><td>Low dose Naltrexon</td></tr>
<tr><td>Paxlovid</td></tr>
<tr><td>Beta blockers for postural tachycardia syndrome (POTS)</td></tr>
<tr><td>Modafinil</td></tr>
<tr><td>Methylphenidate</td></tr>
<tr><td>Fampiridine</td></tr>
<tr><td>Temelimab</td></tr>
<tr><td>Dietary supplements</td></tr>
<tr><td>Pro-/prebiotics</td></tr>
<tr><td>Ginkgo compound</td></tr>
<tr><td>Ginseng root extract</td></tr>
<tr><td>Coenzyme Q</td></tr>
<tr><td>Other – Please specify:</td></tr>
</table>

**Assessment of acute illness**

When was the time of the infection that first led to Post-COVID symptoms? (mm/20yy)

Where you pregnant at the time of the acute COVID infection? (Yes/No)

How many vaccinations have you received before this infection?

| | | | | | | | |
|---|---|---|---|---|---|---|---|
| None | 1 | 2 | 3 | 4 | 5 | 6 | More than 6 |

Which vaccine was used for the the first (second, third, fourth, fifth, sixth) vaccination?

| | | | |
|---|---|---|---|
| Moderna/Spikevax | Pfizer/Biontech | Janssen/Johnson&Johnson | Nuvaxovid/Novavax-NVX-CoV2373 |

Which vaccine was used for the subsequent vaccinations?

Was the diagnosis confirmed? (Yes/No)

How was the diagnosis confirmed?

| | | | |
|---|---|---|---|
| Self-test | PCR | Rapid antigen test at care provider (clinic, test centre) | Antibody test |

What was the highest level of care?

| | | |
|---|---|---|
| Outpatient care | Hospitalisation without intensive care | Hospitalisation with intensive care |

Was there a specific treatment during the acute COVID infection? (Yes/No)

What treatments were used?

| | |
|---|---|
| Oxygen therapy | Invasive |
| | Non-invasive |
| Antivirals | Nirmatrelvic/Ritonavir (Paxlovid) |
| | Remdesivir (Veklury) |
| | Molnupiravir (Lagevrio) |
| | Other – please specify: |
| Neutralising monoclonal antibodies | Sotrovimab (Xevudy) |
| | Tixagevimab/Cligavimab (Evusheld) |
| | Regdanvimab (Regkirona) |
| | Casirivimab/Imdevimab (Ronapreve) |
| | Other - Please specificy: |
| Antithrombotics/Anticoagulation | NMH |
| | UFH |
| Corticosteroids | Dexamethason |
| | Hydrocortison |
| | Prednisolon |
| Antibiotics | Name of active ingredient |
| Other | Please specify |

Was is the first infection? (Yes/No)
It was the x-time infection:
After the initial manifestation of Post-COVID symptoms, have you had a new infection with the corona virus? (Yes/No)
How did the Post-COVID symptoms develop overall after reinfection?

| | | | |
|---|---|---|---|
| Unchanged | Temporarily worse | Persistantly worse | improved |

How many reinfections with SARS-CoV-2 occurred after symptoms of Post-COVID first manifested?
What was the hightet level of care of those reinfections?

| | | |
|---|---|---|
| Outpatient care | Hospitalisation without intensive care | Hospitalisation with intensive care |

**Post-COVID Assessment**

<u>Assessment of acute symtoms</u>

<table>
<tr><th>Symptom (Yes/No)</th><th>Specification (Yes/No)</th><th>Occurance</th><th>Temporal relationship</th></tr>
<tr><td rowspan="2">• Chest Pain<br>• Shortness of breath</td><td rowspan="2"></td><td>At rest</td><td>What is the temporal relationshp between the symptom under strain and the infection?<br>• Pre-existing and of the same intensity<br>• Pre-existing and of higher intensity after the infection<br>• Newly developed after infection</td></tr>
<tr><td>Under strain</td><td>What is the temporal relationship between the symptom at rest and the infection?<br>• Pre-existing and of the same intensity<br>• Pre-existing and of higher intensity after the infection<br>• Newly developed after infection</td></tr>
<tr><td>• Cough<br>• Fatigue<br>• Dizziness/balance impairment<br>• Post Exertional Malaise (PEM)<br>• Palpitations<br>• Orthostatic intolerance<br>• Sleep disturbance<br>• Brain Fog<br>• Headaches</td><td></td><td></td><td>What is the temporal relationship between the symptom and the infection?<br>• Pre-existing and of the same intensity<br>• Pre-existing and of higher intensity after the infection<br>• Newly developed after infection</td></tr>
</table>

| | | | |
|---|---|---|---|
| • Muscle Aches<br>• Muscle Weakness<br>• Skin rash with urticaria<br>• Night Sweats<br>• Tingling/numbness (neurological) | | | |
| Performance intolerance | • Mental<br>• Physical | | |
| Signs of depression | • Less interest or joy regarding activities<br>• Feelings of hopelessness, dejection, or melancholy | | |
| Anxiety/panic attack | • Anxiety<br>• Panic attacks | | |
| Forgetfullness/difficulty concentrating | • Forgetfullness<br>• Difficulty concentrating | | |
| Joint pain/swelling of the joints | • Joint pain<br>• Swelling of the joints | | |
| Impaired sense of smell and/or taste | • Impaired sense of smell<br>• Impaired sense of smell | | |
| Gastrointestinal problems | • Nausea<br>• Emesis<br>• Diarrhea | | |
| Other symptoms | Please specify | | |
| Weight loss | How much (in kg)? | | In what time frame occurred the weight loss (in months)? |

**Medical history and sociobiographical parameter**

<u>Comorbidities at first consultation</u>

| Category of disease | Disease | Additional information | Answer categories |
|---|---|---|---|
| Neoplasia | • Colon<br>• Mammaries<br>• Bronchi<br>• Lymphoma<br>Other – Please specify | • Yes, currently active<br>• Past history<br>• Unknown<br>no | • Yes/No<br>• When was the diagnosis made?<br>○ Preexisting<br>○ Newly diagnosed after infection |
| Cardiovascular disease | • Coronary heart disease<br>• Hypertension<br>• Myocardian infarction<br>• Atrial fibrillation<br>• Deep vein thrombosis<br>• Pulmonary embolism<br>• Hear valce defects<br>• Other – Please specify | | |
| Endocrinological disease | • Type I diabetes<br>• Type II diabetes<br>• Hyperthyroidism<br>• Hypothyroidism<br>• Other – Please specify | | |
| Lung disease | • Asthma<br>• Chronic obstructive pulmonary disease<br>• Obstructive sleep apnea syndrome<br>• Pulmonary fibrosis | | |

| | |
|---|---|
| | • Pulmonary hypertension<br>• Other – Please specify: |
| Psychiatric ilness | • Depression (unipolar/bipolar)<br>• Anxiety disorder<br>• Personality disorder<br>• Delusions/delusional disorder<br>• Other – Please specify: |
| Neurological disorder | • Headaches (migraine/tension headaches)<br>• Multiple sclerosis<br>• Past stroke (cerebrovascular insult/transient ischemic attack)<br>• Other – Please specify: |
| Kidney disease | • Hypertensive nephropathy<br>• Diabetic nephropathy<br>• Immunoglobulin A nephritis (morbus Berger)<br>• Other – Please specify: |
| Immunological disease | • Systemic lupus erythematosus (SLE)<br>• Systemic sclerosis<br>• Polymyositis and dermatomyositis<br>• Sjogren’s syndrome<br>• Mixed collagenoses<br>• Sarcoidosis<br>• Immunoglobulin deficiency<br>• Primary immunodeficiency (except immunoglobulin deficiency)<br>• ANCA-associated small vessel vasculitis<br>• Large cell vasculitis with/without polymyalgia rheumatica<br>• Other – Please specify: |
| Gastrointestinal diseae | • Chronic inflammation of the bowels (Crohn’s disease, ulcerative colitis)<br>• Irritable bowel syndrome (IBS)<br>• Chronic liver disease (cirrhosis, alcoholic steatohepatitis, non-alcoholic steatohepatitis)<br>• Other – Please specify: |
| Chronic infectious disease | • HIV<br>• Hepatitis B<br>• Hepatitis C<br>• Hepatitis D<br>• Hepatitis E<br>• Tuberculosis<br>• Other – Please specify: |
| Allergies/Atopy | • Hay Fever<br>• Allergic asthma<br>• Urticaria<br>• Other – Please specify: |
| Obesity | |

<u>Medication at the time of PCC diagnosis until follow-up</u>
(drug, generic name, newly prescribed in consultation)

- Blood pressure medication:
- Diuretics:
- Anticoagulants (OAC, NOAC, TCAH):
- antiplatelet agents/thrombocyte aggregation inhibitors:

- Statins:
- Antidepressents/anxyiolytics:
- Analgesics:
- Proton pump inhibitors (PPI):
- Inhalants:
- Insulin/Antidiabetics:
- Thyroid medications:
- Immunomodulating drugs (intravenous immunoglobulines, monoclonal antibodies, other immunosuppressants):
- Antihistamines:
- Vitamins/Supplements:
- Other:

<u>Noxious agents</u>

*Nicotine (Yes/No)*

- Pack years:
- No nicotine:
  - Non-smoker
  - Past history of smoking:
    - Pack years:

*Alcohol (Yes/No)*

| | | | | | | | |
|---|---|---|---|---|---|---|---|
| <1/week | 1/week | 2/week | 3/week | 4/week | 5/week | 6/week | Daily |

Past history of alcohol abuse (Yes/No)

*Other drugs (Yes/No)*

- Which?
- Past history of other drugs? Yes/No
- Which other drugs?Yes/No

<u>Pregnancy</u>

- Are you currently pregnant? Yes/No

<u>Social parameters</u>

*Living arrangements*

| | | | |
|---|---|---|---|
| Single household | Multi-person household | Assisted living | Other: Which other living arrangements? |

*Marital status*

| | | | | | |
|---|---|---|---|---|---|
| Unmarried | Married/committed relationship | Divorced | Living separated | Widowed | Other: Which other marital status? |

*Children (Yes/No)*

- Number of children:

*Highest level of education*

- Not (yet) finished secondary education
- finished secondary education
- Apprenticeship/Trade school
- University degree
- Other: Which other level of education?

*What is your current (main) employment status?*

| | | | | | |
|---|---|---|---|---|---|
| Employed | Self-employed | Currently seeking employment | Stay-at-home | Retired | Other |

*Area of occupation*

- Public Relations/Communication
- IT
- Commerce/Trade
- Education
- Media
- Social affairs
- Health/Health Care
- (Business) Consulting
- Logistics
- Project management

- Academia
- Marketing
- Craftmanship
- Production
- Administration
- Electronics
- Software Development
- Assistant
- Coaching and Development
- Industry sector
- Customer Service
- Other

*Which pension?*

| | |
|---|---|
| Retirement pension | Invalidity pension – Basis: |

*Did the patient work prior to the onset of current symptoms? (Yes/No)*

- Was there a change of working quota because of PCC?

| | | |
|---|---|---|
| Yes | No | Unclear |

- Change of quota:

| | | |
|---|---|---|
| Higher quota | Reduced quota | Unclear |

- Are there any special stress factors at work?
  - No
  - Predominantly seated tasks
  - Heavy manual labor (construction, logistics, health care/nursing)
  - Lifting and carrying heavy loads (construction, logistics, health care/nursing)
  - Physical, chemical, biological factors
  - Shift work
  - High time pressure
  - Stressfully high work load
  - Other
- Monotonous and/or demanding work load (e.g., cashier): Yes/No

**Clinical examination and laboratory parameters**

Clinical examination

| | |
|---|---|
| Height (in cm): | Weight (in kg): |
| BMI: | Pulse (BPM) |
| Blood pressure<br>Dystolic/Systolic: __ / __ | Oxygen saturation<br>with room air at rest (in %): __ % |

Laboratory parameters

| | | |
|---|---|---|
| Leukocytes (G/l) | Hemoglobin (g/l) | Bilirubin (µmol/l) |
| Neutrophile (G/l) | Alanine Transaminase (U/l) | Mean cellular volume (fl) |
| Lymphocytes (G/l) | Platelet count (G/l) | Albumin (g/l) |
| Monocytes (G/l) | C-reactive protein (mg/l) | Alkaline phosphatase (U/l) |
| Eosinophile (G/l) | Creatinine kinase (U/l) | Cortisole (nmol/l) |
| Basophile (G/l) | Glycated hemoglobin (HbA1c; %) | Ferritin (µg/l) |
| Sodium (mmol/l) | Potassium (mmol/l) | Creatinine (µmol/l) |
| Free thyroxine (T4; pmol/l) | B12 vitamin (pmol/l) | Phosphate (mmol/l) |
| Anti-nuclear antibodies (titer) | Thyroid stimulating hormone (mIU/l) | Erythrocyte sedimentation rate (mm/h) |
| Calcium (Albumin corrected; mmol/l) | Calcium (Albumin corrected; mmol/l) | |
